# Reductions in 2020 US life expectancy due to COVID-19 and the disproportionate impact on the Black and Latino populations

**DOI:** 10.1101/2020.07.12.20148387

**Authors:** Theresa Andrasfay, Noreen Goldman

**Author notes:** Corresponding Author: Theresa Andrasfay.

## Abstract

COVID-19 has resulted in a staggering death toll in the US: over 215,000 by mid-October 2020, according to the Centers for Disease Control and Prevention. Black and Latino Americans have experienced a disproportionate burden of COVID-19 morbidity and mortality, reflecting persistent structural inequalities that increase risk of exposure to COVID-19 and mortality risk for those infected. We estimate life expectancy at birth and at age 65 for 2020, for the total US population and by race and ethnicity, using four scenarios of deaths – one in which the COVID-19 pandemic had not occurred and three including COVID-19 mortality projections produced by the Institute for Health Metrics and Evaluation. Our medium estimate indicates a reduction in US life expectancy at birth of 1.13 years to 77.48 years, lower than any year since 2003. We also project a 0.87-year reduction in life expectancy at age 65. The Black and Latino populations are estimated to experience declines in life expectancy at birth of 2.10 and 3.05 years, respectively, both of which are several times the 0.68-year reduction for whites. These projections imply an increase of nearly 40% in the Black-white life expectancy gap, from 3.6 to over five years, thereby eliminating progress made in reducing this differential since 2006. Latinos, who have consistently experienced lower mortality than whites (a phenomenon known as the Latino or Hispanic paradox), would see their more than three-year survival advantage reduced to less than one year.

## Introduction

The number of deaths from COVID-19 in the US is staggering: As of mid-October 2020, more than 215,000 COVID-19 deaths had occurred and over 100,000 additional deaths were projected by the end of year (1, 2). An important but as of yet unanswered question concerns the impact of this exceptional number of deaths on life expectancy for the entire nation as well as the consequences for marginalized groups. Despite concerns about inadequate testing and unreliable data, there is convincing evidence that Black and Latino^*^ Americans experience a disproportionate burden of COVID-19 morbidity and mortality. Of the deaths for which race and ethnicity have been reported to the National Center for Health Statistics, 21% were Black and 22% Latino (3).^†^ A plethora of factors likely contribute to these disparities, many of which reflect enduring structural inequalities for the Black and Latino populations that increase both risk of exposure to COVID-19 and risk of death for those infected. Taken together, the high death toll and the racial and ethnic inequities in COVID-19 mortality suggest that COVID-19 will have a major impact on 2020 life expectancy, especially for the Black and Latino populations.

Life expectancy, a frequently used metric of population health that is typically measured as of birth,^‡^ is an informative tool for examining the differential impact of COVID-19 on survival as it is unaffected by the age distribution of the underlying populations. In contrast, for example, comparisons of overall (crude) death rates or proportions of deaths by race and ethnicity are biased by the fact that the Black and Latino populations in the US are younger than the white population and, all else being equal, would have fewer deaths (4, 5).

In the period preceding the COVID-19 pandemic, annual improvements in US life expectancy had been small – e.g., an increase from 76.8 to 78.9 or an average annual increase of 0.15 years between 2000 and 2014 – but overall life expectancy has rarely declined^§^ (6). The recent declines that have taken place have attracted enormous attention from researchers and the media. Annual reductions of 0.1 year for each of three consecutive years (2015, 2016, and 2017) (7–9), attributed partly to increases in “deaths of despair” (6), made repeated headlines as the longest period of decrease since the 1918 influenza pandemic. Conversely, a 0.1-year recovery in life expectancy in 2018 was greeted with substantial relief (10).

Black Americans have consistently had lower life expectancy than whites, but relative gains in life expectancy over the past two decades have been greater in the Black population than among whites, thereby narrowing the Black mortality disadvantage (11, 12). In contrast, because Latinos have consistently had higher life expectancy than whites, a phenomenon referred to as the Latino or Hispanic epidemiological paradox, larger improvements in life expectancy in the Latino population widened the gap between whites and Latinos in recent years, further increasing the Latino mortality advantage (13).

The COVID-19 pandemic has the potential to bring about a greater decline in annual life expectancy than the US has experienced in many years, perhaps since the 1918 influenza pandemic, which is estimated to have reduced US life expectancy between 7 and 12 years (14).^**^

However, the concentration of COVID-19 deaths among the elderly, in contrast to the preponderance of young adult deaths in the previous influenza pandemic (15), may reduce the impact of COVID-19 on life expectancy at birth. The exceptionally high COVID-19 death rates borne by Black and Latino individuals are likely to bring about larger reductions in life expectancy for these populations than for whites, but the pressing question is by how much. In the present analysis, we address this issue by using ongoing data collection of COVID-19 deaths and projections of future deaths under different policy scenarios to estimate how COVID-19 mortality will affect life expectancy at birth and at age 65 for the total population as well as separately by race and ethnicity. We also assess the implications of these projections for the Black disadvantage and the Latino advantage relative to whites.

## Results

We present estimated life expectancy values under four projection scenarios in Table 1. These include one in which the COVID-19 pandemic had not occurred and three projections for the cumulative number of COVID-19 deaths through December 31, 2020 produced by the Institute for Health Metrics and Evaluation (IHME): 1) approximately 321,000 deaths under the current projection scenario, which is the medium scenario issued by IHME; 2) approximately 348,000 deaths under a higher mortality scenario assuming continued easing of mandates; and 3) approximately 276,000 deaths under a lower mortality scenario assuming universal mask usage in the population (1). We estimate that US life expectancy at birth would have been 78.61 years in 2020 had the COVID-19 pandemic not occurred, but all three mortality scenarios imply huge reductions in life expectancy at birth for the US in 2020. The medium scenario would bring about a decline of 1.13 years, whereas the higher and lower mortality scenarios project declines of 1.22 years and 0.98 years, respectively. Life expectancy at age 65, which is estimated to have been 19.40 years in the absence of COVID-19, is projected to decline by 0.87 years under the medium scenario, 0.94 years under the higher mortality scenario, and 0.75 years under the lower mortality scenario.

**Table 1.** Life expectancy projections for the United States in 2020 by race and ethnicity under different COVID-19 mortality scenarios

| Race and ethnicity | Total population |  | Non-Latino white |  | Non-Latino Black |  | Latino |  |
| --- | --- | --- | --- | --- | --- | --- | --- | --- |
|  | Life expectancy |  | Life expectancy |  | Life expectancy |  | Life expectancy |  |
|  | At birth (e<sub>0</sub>) | At age 65 (e<sub>65</sub>) | At birth (e<sub>0</sub>) | At age 65 (e<sub>65</sub>) | At birth (e<sub>0</sub>) | At age 65 (e<sub>65</sub>) | At birth (e<sub>0</sub>) | At age 65 (e<sub>65</sub>) |
| **Absent COVID-19 (referent)** | 78.61 | 19.40 | 78.52 | 19.32 | 74.88 | 18.09 | 81.82 | 21.44 |
| **IHME current projection (medium scenario)** | 77.48 | 18.53 | 77.84 | 18.69 | 72.78 | 16.36 | 78.77 | 19.20 |
| **Difference (years)** | -1.13 | -0.87 | -0.68 | -0.63 | -2.10 | -1.73 | -3.05 | -2.24 |
| **IHME mandates easing scenario (higher mortality scenario)** | 77.39 | 18.46 | 77.79 | 18.64 | 72.62 | 16.23 | 78.54 | 19.03 |
| **Difference (years)** | -1.22 | -0.94 | -0.73 | -0.68 | -2.26 | -1.86 | -3.28 | -2.41 |
| **IHME universal masks scenario (lower mortality scenario)** | 77.63 | 18.65 | 77.93 | 18.78 | 73.05 | 16.58 | 79.16 | 19.48 |
| **Difference (years)** | -0.98 | -0.75 | -0.59 | -0.54 | -1.83 | -1.51 | -2.66 | -1.96 |

Source: Authors' calculations of life expectancy projections in 2020. Difference refers to the difference between the projection and the absent COVID-19 scenario. Calculations are based on IHME projections that were updated October 9, 2020.

Estimated life expectancy values in the absence of COVID-19 compared with those under the medium scenario are displayed in Figure 1, which underscores the much larger reductions in life expectancy anticipated for the Black and Latino populations than for whites or the total US. Under the medium scenario, white life expectancy is projected to decline by 0.68 years while the corresponding declines for Black and Latino life expectancies are 2.10 years and 3.05 years, respectively. We also observe racial and ethnic disparities in the projected impact of COVID-19 on remaining life expectancy at age 65, which is estimated to decline by 0.63 years for the white population, 1.73 years for the Black population, and 2.24 years for the Latino population.

**Figure 1.**
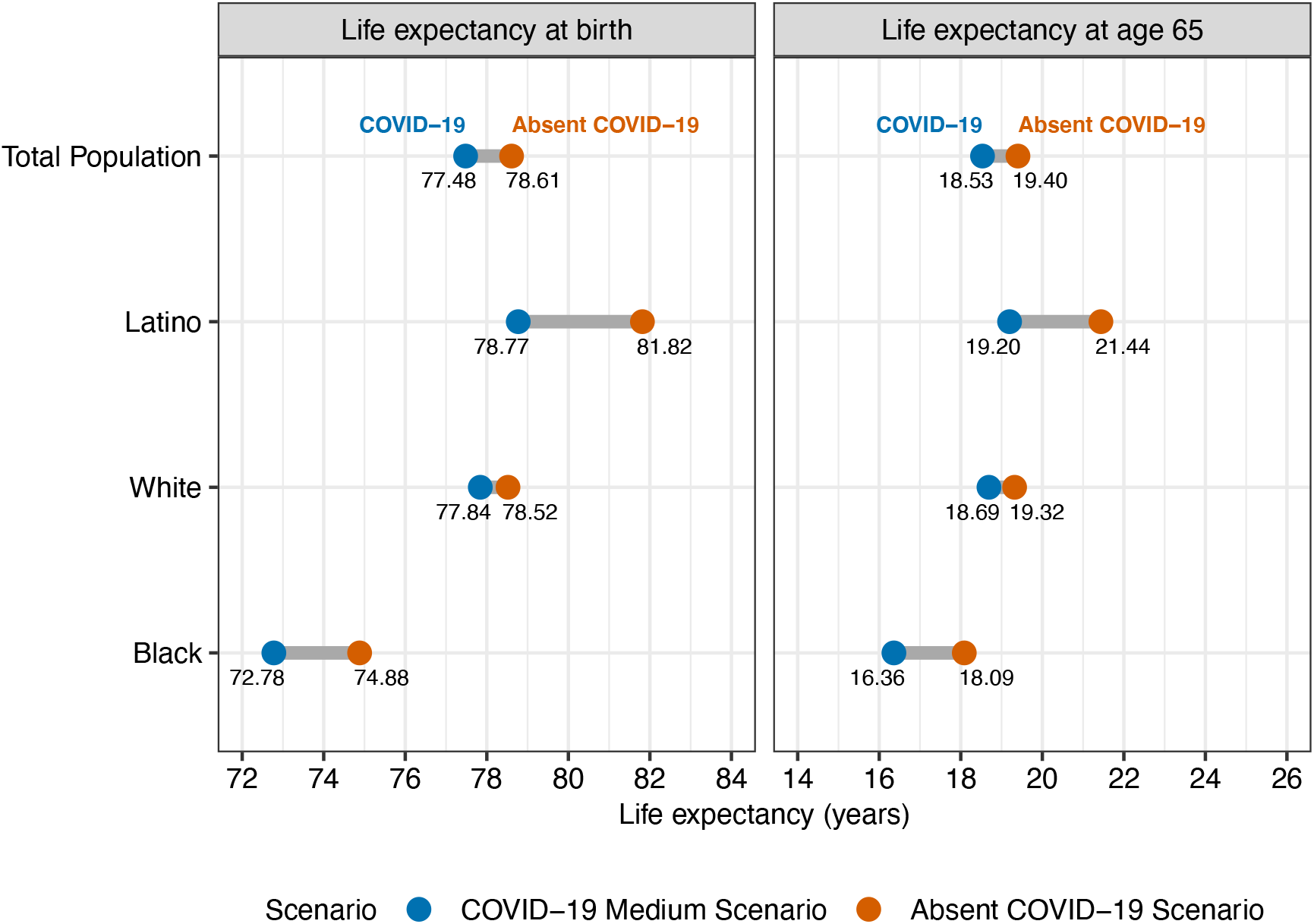
Life expectancy projections for 2020 by race and ethnicity in the absence of COVID-19 and under the medium scenario. The COVID-19 medium scenario is based on the IHME current projection scenario (October 9, 2020 update).

For whites, the projected effect of COVID-19 is similar for life expectancy at birth and at age 65; however, for the Black and Latino populations, the effect on life expectancy at birth is notably larger than for life expectancy at age 65, reflecting the higher burden of COVID-19 mortality at younger ages among these groups. These racial and ethnic differences are also revealed by identifying the age groups showing the largest proportional increases in mortality rates in the presence of COVID-19: 75-85 and 85 and over in the white population, 65-75 and 75-85 in the Black population, and 55-65 and 65-75 in the Latino population (*Supplementary material*).

Notable racial and ethnic disparities in life expectancy declines are projected under both of the alternative COVID-19 mortality scenarios. Under the higher mortality scenario, life expectancy is projected to be 0.73 years lower for the white population, 2.26 years lower for the Black population, and 3.28 years lower for the Latino population. Under the lower mortality scenario, life expectancy at birth is projected to decline by 0.59 years for the white population, 1.83 years for the Black population, and 2.66 years for the Latino population.

To put these projected life expectancy declines in perspective, trends in life expectancy at birth from 1980-2020 are presented in Figure 2 by race and ethnicity. The projected decline in life expectancy due to COVID-19 for the US, under the medium scenario, is larger than any other single-year decline during this time period and would return US life expectancy to a value (77.48 years) lower than that observed in any year since 2003.

**Figure 2.**
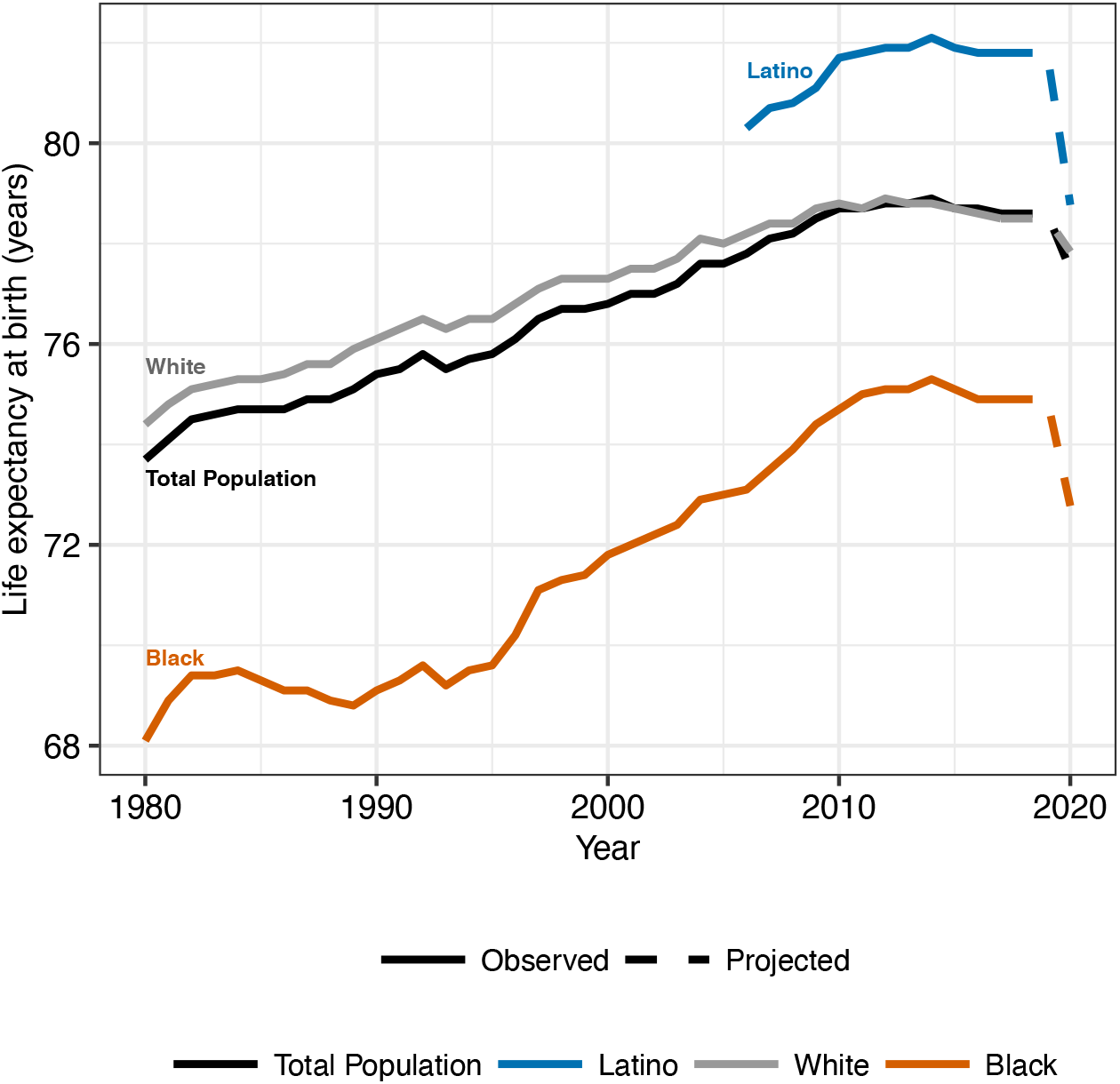
Trends in life expectancy at birth by race and ethnicity: 1980-2020. Note: The data for the Black and white populations prior to 2006 include Latinos; data for these groups from 2006 onward are for the non-Latino Black and non-Latino white populations. The projections for 2020 are based on the IHME current projection scenario (October 9, 2020 update).

To illustrate the effect of COVID-19 on the racial and ethnic differences in life expectancy at birth, Figure 3 displays the difference in life expectancy for the Black and Latino populations relative to the white population. Following a widening in the late 1980s, the Black-white life expectancy gap has narrowed from 7.1 years in 1993 to 3.6 years in 2017. Under the medium scenario, this Black disadvantage is projected to widen to 5.06 years in 2020, which is the size of the gap in 2006. In contrast, since 2006 (the earliest year for which Latinos are separately identified in Vital Statistics), the Latino population has experienced a growing life expectancy advantage relative to whites, increasing from an advantage of 2.1 years in 2006 to 3.3 years in 2017. Under the medium scenario, this advantage is estimated to decline to 0.93 years in 2020, smaller than ever recorded nationally. The Black-white gap would widen to 5.17 years under the higher mortality scenario and 4.88 years under the lower mortality scenario. The Latino advantage would narrow to 0.75 years under the higher mortality scenario and 1.23 years under the lower mortality scenario (see Table 1 for projected life expectancy values under alternative scenarios).

**Figure 3.**
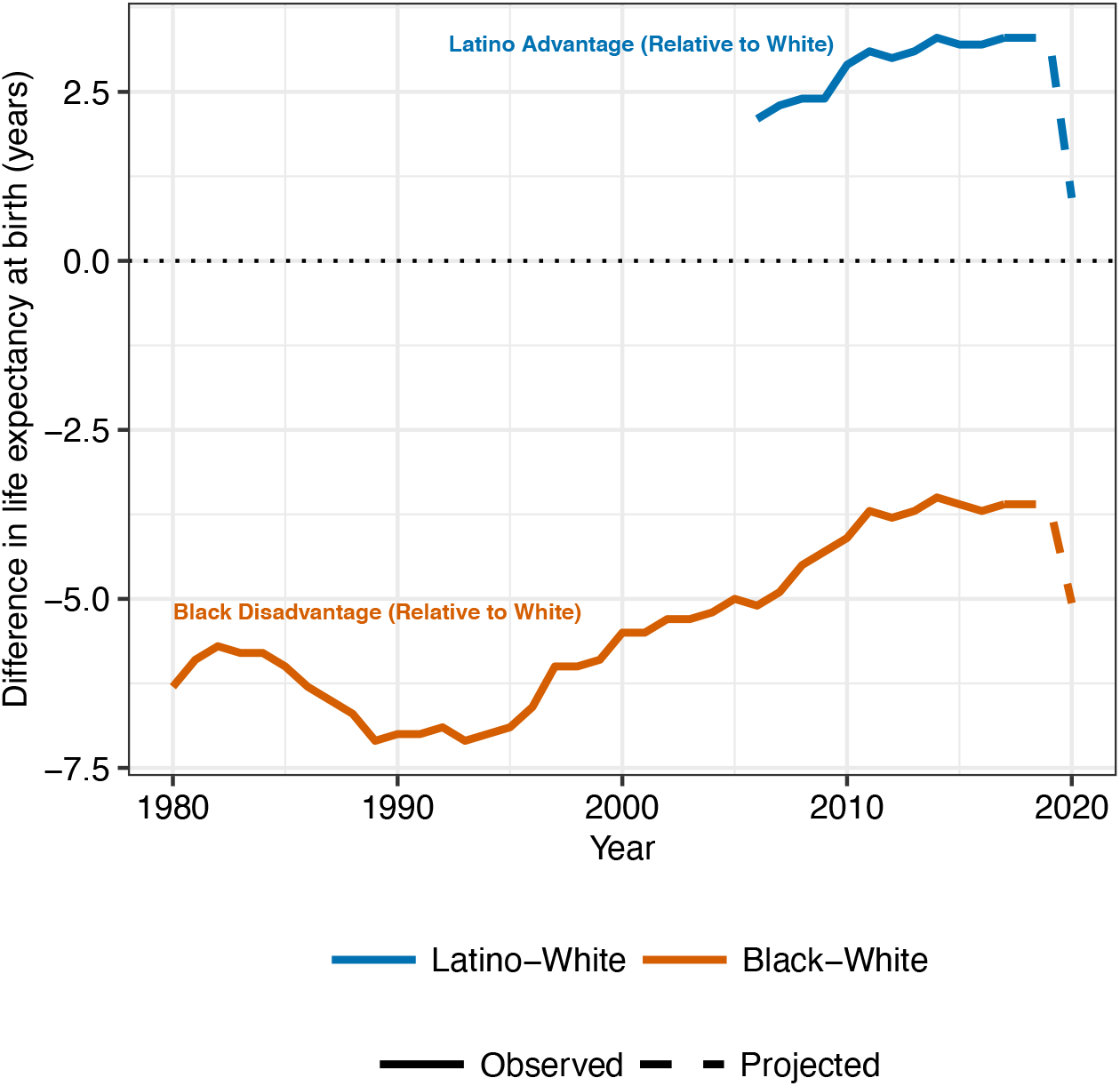
Trends in racial and ethnic differences in life expectancy at birth: 1980-2020. Note: The data for the Black and white populations prior to 2006 include Latinos; data for these groups from 2006 onward are for the non-Latino Black and non-Latino white populations. The projections for 2020 are based on the IHME current projection scenario (October 9, 2020 update).

## Discussion

Our findings reveal that, according to the medium scenario of COVID-19 mortality, the pandemic is projected to result in an enormous decline in 2020 US period life expectancy of 1.13 years. This estimate is similar to that calculated by Heuveline and Tzen in August 2020 (16). This impact is about ten times as large as the worrisome annual decreases several years ago that were attributed largely to drug overdoses, other external causes, and respiratory and cardiovascular diseases (6, 17). The US reduction in 2020 life expectancy is projected to exceed that of most other high-income countries (16), indicating that the US – which already had a life expectancy below that of all other high-income developed nations prior to the pandemic (17) – will see its life expectancy fall even further behind its peers. Although the majority of COVID-19 deaths in 2020 have likely already occurred, our projections under different mortality scenarios underscore the potential for policy interventions and public behavior to either mitigate or exacerbate the ultimate effect of COVID-19 on 2020 life expectancy. In particular, estimated declines in life expectancy for all racial and ethnic groups under the lower mortality universal mask scenario are reduced by over 12%, in contrast to an increase of over 7% under the higher mortality scenario that assumes continued easing of mandates.

In addition, we estimate that the pandemic will result in reductions in life expectancy for the Black (2.10 years) and Latino (3.05 years) populations that are three to four times as large as the reduction for whites (0.68 years). The greater toll for the Black and Latino populations arises because of both higher COVID-19 mortality rates and greater susceptibility to COVID-19 at younger ages among these groups compared with whites. In both of the alternative mortality scenarios, the disparities would remain immense.

Potential explanations for the disproportionate burden of deaths among Black and Latino individuals reflect underlying social disparities that have been documented for decades and amplified during the current pandemic. These groups are more likely than whites to hold low paying jobs with little autonomy, often in industries that have suffered the largest job losses during the pandemic, creating exceptionally high unemployment rates for both groups and likely loss of health insurance (18, 19). Many of those who retained their jobs in essential industries, such as health care, food retail, and meatpacking, face high exposure to viral transmission (20, 21). The incompatibility of their jobs with remote working, combined with low savings, likely compelled many Black and Latino workers to continue to risk exposure to COVID-19 (22, 23). Undocumented Latinos have been facing the further disadvantage of being ineligible for government unemployment benefits. Living situations provide additional exposure to infection for these groups since they are more likely than whites to reside in crowded, multi-generational housing units (24, 25) and densely populated areas (26, 27) and to rely on public transportation (23). Lower rates of health insurance, particularly among undocumented Latinos (28, 29), and generally poorer quality of care inside and outside of hospitals compared with whites (30, 31) may have increased the risk that COVID-19 infection among Black and Latino individuals became fatal.

The risk of COVID-19 mortality is also heightened by several chronic conditions that are prevalent at older ages, such as hypertension, obesity, diabetes, cancer, and heart disease (32–34). Black adults generally have higher rates of these co-morbidities, as well as higher death rates from these conditions, than whites or Latinos, often developing these conditions at much younger ages (33, 35–37). In contrast, Latinos report lower rates of cancer and heart disease (33, 38) compared with both white and Black adults, although they experience relatively high rates of both obesity and diabetes (33, 35, 36, 38).

One of many very distressing consequences of the COVID-19 pandemic is an estimated 39% increase in the Black-white life expectancy gap, reversing progress made in reducing this disparity since 2006. Since the US government began compiling relatively complete death registration data nationally in 1929, mortality risks documented for the Black population have exceeded those for whites, with the Black-white life expectancy gap peaking at 13.3 years in 1930 (39). The Black life expectancy disadvantage has generally held steady or declined over the latter half of the 20^th^ century with the notable exception of the 1980s, when the gap widened partly as a result of increased rates of HIV and homicide among Black men (11, 39, 40). However, since the early 1990s, death rates among Black individuals have declined more rapidly than for whites, largely due to a reduction in these causes as well as declines in heart disease and some types of cancer in the Black population and disproportionate increases in unintentional poisonings among whites (11, 12, 41), cutting the 7-year gap in life expectancy at birth in 1990 to almost half in 2017. Our medium estimate of the impact of COVID-19 mortality suggests that the Black-white gap will increase again to over five years in 2020. An alternative perspective on the Black-white life expectancy gap in 2020 is provided by Wrigley-Field (2020), who estimates that over 600,000 excess white deaths would need to occur in 2020 to lower white life expectancy to the highest life expectancy values ever experienced by the Black population (42).

The findings for Latinos reveal an interesting conundrum in that Latinos have experienced better survival than white and especially Black individuals throughout the period of data availability. This “paradox,” so-called because Latinos experience higher longevity than whites despite having substantially lower education, income, wealth and access to health care, is most pronounced among immigrants and has been shown to result partly from relatively low rates of smoking in past decades as well as selective migration, particularly emigration from the US of Latinos in poor health (13, 43–45). The generally good health of Latinos, which all else being equal should have protected them from COVID-19 infection and fatalities, has laid bare the risks associated with social and economic disadvantage. Our estimates indicate an unprecedented mortality increase for Latinos, exceeding that for the Black population, that would eliminate over 70% of the previous Latino advantage relative to whites.

This analysis contains several limitations. These calculations rest on several assumptions (*Materials and Methods*), including that the age, racial, and ethnic distributions of future COVID-19 deaths will be equivalent to the distributions reported for current deaths. Data on deaths from COVID-19 are likely incomplete for several reasons, including attribution of COVID-19 deaths to other causes and incomplete and inaccurate recording of age, race, and ethnicity of these deaths.

We also assume that individuals who do not die from COVID-19 experience the mortality conditions observed in 2017. This assumption does not allow us to include the impact of excess deaths from other causes that may have been related to the pandemic, such as deaths that could have been prevented had individuals not delayed or forgone medical care because of fear of contracting COVID-19, lost their health insurance, or faced other disruptions produced by the pandemic. Estimates of excess mortality suggest that deaths attributed to COVID-19 account for only two-thirds to three-fourths of all excess deaths in the US (46, 47). The impact of this underestimate of deaths in 2020 on life expectancy may be counteracted by harvesting – i.e., the notion that COVID-19 might disproportionately affect frail individuals with severe health conditions who would thus be likely to die imminently from other causes in the absence of COVID-19 (48) – but there is no evidence that harvesting has played a notable role in COVID-19 mortality.

Our predictions are based on IHME scenarios that were issued on October 9, 2020. At this time, the situation is rapidly evolving, and future deaths will depend on many factors, including government policies, human behavior, and scientific advances. Thus, our projections should be interpreted as our best estimates of what will happen to life expectancy in 2020, rather than a definitive forecast.

Life expectancy does not describe the lived or anticipated experience of any actual cohort or group of people. Rather, life expectancy at a given age (e.g., birth or age 65) is an estimate of how long a population or a group of individuals would live if they were to experience the sequence of age-specific death rates observed in a given period, such as a calendar year, starting at birth or age 65 and continuing through their remaining lifetime. In light of the expectation that the COVID-19 pandemic will subside with the development of vaccines, treatments, and long-term behavioral changes to reduce exposure, no cohort may ever experience a reduction in life expectancy of the magnitude attributed to COVID-19 in 2020. At the same time, however, a rapid return to pre-COVID-19 life expectancy is unlikely due to the anticipated continued presence of the SARS-COV-2 virus, long-term detrimental health impacts for those who recovered from the virus, deaths from other health conditions that were precipitated by COVID-19, and social and economic losses resulting from the pandemic (49).

## Materials and Methods

### Data

We draw on data from multiple sources to calculate projections of period life expectancy in 2020 for the entire population and the non-Latino white, non-Latino Black, and Latino populations in the United States, excluding Puerto Rico. Counts of deaths involving COVID-19 by race, ethnicity and age group are obtained from the National Center for Health Statistics (NCHS) for deaths through October 3, 2020 (3).^††^ The projected total numbers of COVID-19 deaths by December 31, 2020 under three policy and behavioral scenarios are obtained from the Institute for Health Metrics and Evaluation (IHME); the projections used in our calculations were updated October 9, 2020 (1).

Mid-year population counts by race, ethnicity, and age are obtained from the US Census Bureau’s estimates for 2019, the most recent year for which these population counts are available (50). Estimates of age-specific mortality rates and other life table measures are taken from life tables published by the National Vital Statistics System for the year 2017, the most recent year for which these life tables are available; separate life tables are obtained for each of the racial and ethnic groups in our study (39). We assume that the 2020 population counts are equivalent to the 2019 population estimates and that the mortality rates for 2020 in the absence of COVID-19 would be equivalent to those observed in 2017.

### Methods

We estimate 2020 life expectancy under four scenarios, including one in which the COVID-19 pandemic had not occurred and three IHME projections based on different levels of COVID-19 mortality: the current projection scenario, which we refer to as the medium scenario, the mandates easing scenario, which we refer to as the higher mortality scenario, and the universal mask scenario, which we refer to as the lower mortality scenario. These scenarios are further described in the supplementary material. Due to differences in state reporting and incomplete adherence to reporting guidelines, race, ethnicity and age are not available for all deaths involving COVID-19. At present, there are no published projections for the total number of US COVID-19 deaths in 2020 by race and ethnicity. To address these data limitations, we assume that the total projected national number of deaths from COVID-19 will have the same racial, ethnic, and age distribution as the COVID-19 provisional death counts published by NCHS for deaths through October 3, 2020 for which race, ethnicity, and age are available.

Our calculation comprises several steps. First, we calculate the expected number of deaths in 2020 in the absence of any COVID-19 mortality by applying the 2017 age-specific mortality rates to the 2019 population counts by age group. Second, we estimate the projected total number of deaths in 2020 by adding the total projected number of deaths from COVID-19 by age group to the expected number of deaths from other causes within each age group. To calculate the expected number of deaths from causes other than COVID-19, we assume that all individuals who do not die of COVID-19 in 2020 are subject to the 2017 age-specific mortality rates. Third, we calculate the age-specific ratio of expected number of deaths in the absence of COVID-19 mortality to the projected total number of deaths in 2020. Using this ratio and standard life table techniques, we then treat the 2017 life table as a cause-deleted life table in which COVID-19 is the cause of death that has been eliminated and recover an all-cause life table that includes deaths from COVID-19; this approach has been used to estimate COVID-19 life expectancy reductions across countries (16). These calculations are done separately for each racial and ethnic group and projection scenario. Additional details on these calculations are provided in the supplementary material.

## Supporting information

Life Expectancy Calculations

## Data Availability

The data used in this study are all publicly available through the provided links.

https://data.cdc.gov/NCHS/Deaths-involving-coronavirus-disease-2019-COVID-19/ks3g-spdg

https://www.cdc.gov/nchs/data/nvsr/nvsr68/nvsr68_07-508.pdf

https://www.census.gov/newsroom/press-kits/2020/population-estimates-detailed.html

https://covid19.healthdata.org/

## Data Availability

The data used in this study are all publicly available through the provided links.

https://data.cdc.gov/NCHS/Deaths-involving-coronavirus-disease-2019-COVID-19/ks3g-spdg

https://www.cdc.gov/nchs/data/nvsr/nvsr68/nvsr68_07-508.pdf

https://www.census.gov/newsroom/press-kits/2020/population-estimates-detailed.html

https://covid19.healthdata.org/

## Data Availability

The data used in this study are all publicly available through the provided links.

https://data.cdc.gov/NCHS/Deaths-involving-coronavirus-disease-2019-COVID-19/ks3g-spdg

https://www.cdc.gov/nchs/data/nvsr/nvsr68/nvsr68_07-508.pdf

https://www.census.gov/newsroom/press-kits/2020/population-estimates-detailed.html

https://covid19.healthdata.org/

## Data Availability

The data used in this study are all publicly available through the provided links.

https://data.cdc.gov/NCHS/Deaths-involving-coronavirus-disease-2019-COVID-19/ks3g-spdg

https://www.cdc.gov/nchs/data/nvsr/nvsr68/nvsr68_07-508.pdf

https://www.census.gov/newsroom/press-kits/2020/population-estimates-detailed.html

https://covid19.healthdata.org/

## Footnotes

* We use the terms “Latino” to refer to individuals with Hispanic or Latino origin, regardless of race, “Black” to refer to non-Hispanic/non-Latino Black or African American, and “white” to non-Hispanic/non-Latino white.

† These percentages are calculated from death certificates with demographic characteristics (age, race, and ethnicity) that have been reported to and processed by the National Center for Health Statistics as of October 7, 2020.

‡ Throughout this paper, life expectancy refers to life expectancy at birth unless specified at another age.

§ Throughout this paper, life expectancy values taken from external publications are reported with one decimal place; our calculations are reported with two decimal places.

** Currently, the largest documented reduction in life expectancy since the 1918 pandemic was a 2.9-year reduction between 1942 and 1943 (NCHS, 2018). NCHS estimates of life expectancy before 1948 used an interpolation method for the intercensal years that relied on a strong assumption about the stationarity of the population age structure (Smith and Bradshaw 2006). Recalculations by Smith and Bradshaw (2006) using annual estimated life tables for these years suggest the reduction in life expectancy between 1942 and 1943 was closer to 0.1 year.

†† These include all deaths for which the underlying cause or a contributing cause of death is ICD-10 code U07.1 (ICD-10 code for COVID-19) regardless of whether the infection was laboratory-confirmed. These are provisional counts from death certificates reported to NCHS that have not been adjusted for delays in reporting or underreporting. Our calculations suggest that the provisional counts by race, ethnicity, and age for deaths through October 3 covered approximately 94% of deaths that were reported by local jurisdictions as of this date.

