## Supplementary Material for "Reductions in 2020 US life expectancy due to COVID-19 and the disproportionate impact on the Black and Latino populations"

### Data Sources

COVID-19 death counts by race, ethnicity[1] and age group are obtained from the National Center for Health Statistics (NCHS) (1). These are provisional counts from death certificates reported to NCHS that have not been adjusted for delays in reporting or underreporting. At the time of writing, this demographic information is available for deaths through October 3, 2020, though NCHS cautions that data for this period are incomplete because of delays in reporting to NCHS. Our calculations suggest that the provisional counts by race, ethnicity, and age for deaths through October 3, 2020 covered approximately 94% of deaths that were reported by local jurisdictions as of this date. These counts include all deaths for which the underlying cause or a contributing cause of death is ICD-10 code U07.1 (ICD-10 code for COVID-19) regardless of whether the infection was laboratory-confirmed.

Total projected numbers of deaths from COVID-19 in the United States are obtained from the Institute for Health Metrics and Evaluation (IHME) (2). The numbers used in the calculations were updated by IHME on October 9, 2020 for deaths through February 1, 2021; we use projected deaths through December 31, 2020. IHME produces estimates for deaths under three scenarios. Additional detail on the methodology and rationale underlying these scenarios are available from IHME, but they are described briefly below:

1. The current projection is considered by IHME to be the most likely scenario. It assumes that each location will continue to ease mandates unless a location exhibits a daily death rate above 8 per million, at which point these mandates will be reinstated for six weeks.
2. The mandates easing scenario assumes that each location will continue to ease mandates regardless of the daily death rate.
3. The universal masks scenario assumes that 95% of the population wears masks whenever they are in public. Like the current projection model, this scenario assumes mandates will continue to ease unless deaths exceed the 8 per million threshold.

Mid-year population counts by race, ethnicity, and age are obtained from the US Census Bureau's estimates for 2019, the most recent year for which these population counts are available (3). We assume that the 2020 population distribution by race, ethnicity, and age is equivalent to the 2019 population distribution.

Estimates of mortality rates and other life table quantities by race and ethnicity and age are taken from the US life tables published by the National Vital Statistics System for the

[1] In the text, we use the term "Latino" to refer to individuals with Latino or Hispanic origin. In this supplementary material and the excel document, we use the term from the original source.

year 2017, the most recent year for which these life tables are available (4). We assume that, in the absence of COVID-19, mortality conditions in 2020 would be equivalent to those observed in 2017.

### Methods

The calculations are provided in the excel document "Life Expectancy Projections-2020-10-15". Each of the excel sheets is described below.

**Sheet 1: Total-2017-Life Table**
This sheet is the life table for the total population in the United States in 2017 taken directly from Table 1 in the National Vital Statistics Reports United States Life Tables, 2017. This life table is unabridged, reporting life table quantities for single-year age intervals from birth to age 100.

**Sheet 2: White-2017-Life Table**
This sheet is the life table for the non-Hispanic white population in the United States in 2017 taken directly from Table 13 in the National Vital Statistics Reports United States Life Tables, 2017. This life table is unabridged, reporting life table quantities for single-year age intervals from birth to age 100.

**Sheet 3: Black-2017-Life Table**
This sheet is the life table for the non-Hispanic Black population in the United States in 2017 taken directly from Table 16 in the National Vital Statistics Reports United States Life Tables, 2017. This life table is unabridged, reporting life table quantities for single-year age intervals from birth to age 100.

**Sheet 4: Hispanic-2017-Life Table**
This sheet is the life table for the Hispanic population in the United States in 2017 taken directly from Table 10 in the National Vital Statistics Reports United States Life Tables, 2017. This life table is unabridged, reporting life table quantities for single-year age intervals from birth to age 100.

**Sheet 5: Life Table-Total-Adjusted**
This sheet aggregates the life table from sheet 1 into wider age intervals that correspond to the age intervals reported for COVID-19 deaths and population counts.
The $l_x$ column of this sheet takes the $l_x$ value for each age x from the original unabridged life table (sheet 1) to obtain the survivorship at age x.
The ${}_nL_x$ column takes the single-year ${}_1L_x$ values from the original unabridged life table (sheet 1) and sums these values from ages x to x + n to obtain the total period person-years lived between ages x and x + n (${}_nL_x$).
The remaining columns, corresponding to the life table values for person-years lived above age x ($T_x$), life expectancy at age x ($e_x$), expected number of deaths between ages x and x + n (${}_nd_x$), probability of surviving between ages x and x + n (${}_np_x$), probability of dying between ages x and x + n (${}_nq_x$), average number of years lived between ages x and x + n by those who died between ages x and x + n (${}_na_x$), and mortality rate between ages x

and x + n ($_nM_x$), are derived from the $l_x$ and $_nL_x$ columns and standard life table relationships (5).

**Sheet 6: Life Table-White-Adjusted**
This sheet aggregates the life table from sheet 2 into wider age intervals that correspond to the age intervals reported for COVID-19 deaths and population counts. See sheet 5 description for more details.

**Sheet 7: Life Table-Black-Adjusted**
This sheet aggregates the life table from sheet 3 into wider age intervals that correspond to the age intervals reported for COVID-19 deaths and population counts. See sheet 5 description for more details.

**Sheet 8: Life Table-Hispanic-Adjusted**
This sheet aggregates the life table from sheet 4 into wider age intervals that correspond to the age intervals reported for COVID-19 deaths and population counts. See sheet 5 description for more details.

**Sheet 9: Death Totals COVID-19**
This sheet lists the number of total COVID-19 deaths in the United States projected by IHME under the various scenarios. Additional columns provide details on the source of these data, the projection period, and the date these projections were updated.

**Sheet 10: Population Counts**
The upper section of this sheet is taken directly from the Census Bureau's 2019 mid-year population estimates by age, race, and ethnicity. The lower section of this sheet aggregates these population counts into wider age intervals to correspond to the age intervals reported for COVID-19 deaths.

**Sheet 11: COVID-19 Deaths Race-Eth-Age**
This sheet lists the number of deaths involving COVID-19 reported to NCHS by race, Hispanic origin, and age group for the United States. These data are from deaths through October 3, 2020.
We then calculate the percentage of all COVID-19 deaths with known race, ethnicity, and age that are in each of these cells. Next, we apply these percentages to the total number of deaths to date and under each of the three projection scenarios to obtain the expected number total of deaths by race, ethnicity, and age. This assumes that the nationwide total COVID-19 deaths will have the same racial, ethnic and age distribution as the deaths reported to the CDC with this information available.

**Sheet 12: Life Expectancy-Main**
Life expectancy calculations for 2020 using death counts for the main scenario, the IHME current projection scenario. The strategy here is to treat the 2017 life table as the cause-deleted life table in which COVID-19 is the cause that has been deleted and then recover the all-cause life table that includes deaths from COVID-19. A cause-deleted life table, also known as an associated single decrement life table, is a hypothetical set of age-

specific mortality rates and other life table quantities that would be expected if a single cause of death were eliminated (5).

To do so, we do the following intermediate calculations for each racial and ethnic group:

1. Calculate estimated number of deaths in 2020 in the absence of COVID-19 (${}_nD_x{}^*$) by applying the 2017 age-specific death rates (${}_nM_x{}^{17}$) to the 2019 population count by age interval (${}_nK_x{}^{19}$).

$$ {}_nD_x{}^{*} = {}_nM_x{}^{17} * {}_nK_x{}^{19} $$

2. Calculate estimated number of deaths in 2020 with COVID-19 by applying the 2017 age-specific death rates (${}_nM_x{}^{17}$) to the 2019 population count by age interval (${}_nK_x{}^{19}$), less the number projected to die from COVID-19 in this age group. Then we add in the total projected COVID-19 deaths by age group.

$$ {}_nD_x{}^{20} = {}_nM_x{}^{17} * \left[ {}_nK_x{}^{19} - {}_nD_x{}^{COV} \right] + {}_nD_x{}^{COV} $$

where ${}_nD_x{}^{20}$ is the estimated number of deaths between ages x and x + n in 2020 and ${}_nD_x{}^{COV}$ is the estimated number of deaths between ages x and x + n from COVID-19 (sheet 11).

3. Calculate the age-specific ratio of deaths in the absence of COVID-19 to deaths in the presence of COVID-19. This is equivalent to the ratio of deaths from a single cause (in this case deaths from all but one cause) to deaths from all causes (${}_nR_x$) from Chiang's method. (6)

$$ {}_nR_x = \frac{{}_nD_x{}^{*}}{{}_nD_x{}^{20}} $$

4. Calculate the inverse of ${}_nR_x$, which simplifies some later calculations and can be interpreted as the ratio of deaths or mortality rates in the presence of COVID-19 to deaths or mortality rates in the absence of COVID-19.
5. Using Chiang's method, which assumes that the force of decrement from cause *i* (or, alternatively, all causes except *i*) is proportional to the force of decrement from all other causes, we can calculate ${}_np_x{}^{20}$, ${}_nq_x{}^{20}$ and ${}_na_x{}^{20}$ in the presence of COVID-19:

$$ {}_np_x{}^{20} = e^{\left(\frac{1}{{}_nR_x} * \log({}_np_x{}^{17})\right)} $$

$$ {}_nq_x{}^{20} = 1 - {}_np_x{}^{20} $$

$$ {}_na_x{}^{20} = n + \frac{1}{{}_nR_x} * \frac{{}_nq_x{}^{17}}{{}_nq_x{}^{20}} * \left( {}_na_x{}^{17} - n \right) $$

$$ {}_\infty a_{85}{}^{20} = {}_\infty a_{85}{}^{17} * {}_nR_x $$

We then complete the rest of the life table for 2020 in the presence of COVID-19 using standard life table relationships (5). This process is separately repeated for all racial and ethnic groups.

**Sheet 13: Life Expectancy-Easing**
This sheet repeats the calculations from sheet 12 using death counts from the mandates easing scenario.

**Sheet 14: Life Expectancy-Mask**
This sheet repeats the calculations from sheet 12 using death counts from the universal mask scenario.

**Sheet 15: Manuscript Table**
This sheet consolidates the results we report as Table 1 in the text. This sheet also calculates changes in the Black-white and Latino-white life expectancy gaps that are reported in the text.
